# Sensitivity of nasopharyngeal, oropharyngeal and nasal washes specimens for SARS-CoV-2 detection in the setting of sampling device shortage

**DOI:** 10.1101/2020.08.01.20166397

**Authors:** Calame Adrien, Mazza Léna, Renzoni Adriana, Kaiser Laurent, Schibler Manuel

**Author notes:** Corresponding author*: Adrien Calame, Laboratory of Virology, Division of Infectious Diseases, Geneva University Hospitals, Rue Gabrielle-Perret-Gentil 4, 1205 Geneva 14, Switzerland. Calame Adrien and Mazza Léna contributed equally to this work. Kaiser Laurent and Schibler Manuel contributed equally to this work. **Author contributions** All authors contributed to the study conception and design. Material preparation and data collection were performed by Calame Adrien and Mazza Léna. Laboratory analyses were performed by Renzoni Adriana. Statistical analyses were performed by Calame Adrien. The first draft of the manuscript was written by Calame Adrien and Mazza Léna. The final version of the manuscript was written and corrected by Calame Adrien, Mazza Léna, Kaiser Laurent and Schibler Manuel. All authors read and approved the final manuscript.

## Abstract

In the context of an unprecedented shortage of nasopharyngeal swabs (NPS) or sample transport media during the coronavirus disease 2019 (COVID-19) crisis, alternative methods for sample collection are needed. To address this need, we validated a cell culture medium as a viral transport medium, and compared the analytical sensitivity of SARS-CoV-2 real-time RT-PCR in nasal wash (NW), oropharyngeal swab (OPS) and NPS specimens. Both the clinical and analytical sensitivity were comparable in these three sample types. OPS and NW specimens may therefore represent suitable alternatives to NPS for SARS-CoV-2 detection.

## Introduction

In the ongoing Coronavirus Disease 2019 (COVID-19) pandemic caused by severe acute respiratory syndrome coronavirus 2 (SARS-CoV-2), a broad testing strategy is crucial to identify infected persons, including less typical clinical presentations of the disease [1]. However, during this pandemic, broad screening is sometimes hampered by equipment and reagent shortages occurring worldwide [2]. Affected items include sampling devices, as well as molecular testing reagents and viral transport medium.

The Human Coronaviruses (HCoVs) have been identified in a variety of specimens, including oropharyngeal, nasopharyngeal, nasal, sputum and bronchial fluid specimens [3, 5]. The detection of the sarbecovirus SARS-CoV-2, which causes COVID-19, by real-time reverse-transcription PCR (rRT-PCR) using a nasopharyngeal specimen is by now the most commonly accepted method and is recommended by the American CDC and others [https://www.cdc.gov/coronavirus/2019-ncov/hcp/clinical-criteria.html]. SARS-CoV-2 can also be found in oropharyngeal, sputum or even saliva specimens [6-10]. The WHO recommends collecting nasopharyngeal swabs (NPS), oropharyngeal swabs (OPS) or nasal wash (NW) specimens from ambulatory patients with COVID-19 disease [https://apps.who.int/iris/handle/10665/331329?locale-attribute=fr&].

In this crisis setting our institution, like many others, risked facing an unprecedented shortage of equipment, including NPS. We therefore tested several procedures in order to validate alternative solutions in house. OPS and NW are the two alternative procedures presented in this article. They are compared to the gold standard, the NPS. Finally, we validated the use of Dulbecco’s Modified Eagle Medium (DMEM) as a transport medium for SARS-CoV-2 specimens.

## Material and methods

### Participants

Eligible participants were ≥ 18 years old and hospitalized in the internal medicine wards at the Geneva University Hospitals, who had a positive SARS-CoV-2 rRT-PCR in a NPS specimen in the preceding one to six days. ICU patients were excluded.

### Oropharyngeal swabs sampling

PCR tubes (CobasTM Roche Reference No. 07976577001-3) were filled with 3ml of DMEM. Nylon flocked NPS (COPAN Reference No.A305CS01) and cotton OPS (VWR Reference No. 300260) were used for sampling. Consecutive OP and then NP samples from 29 patients were obtained in parallel and inserted into DMEM Cobas tubes. The OPS was always performed first. OPS specimens were obtained by swabbing the oropharyngeal posterior wall and turning once in each direction. They were then transferred into the Cobas PCR tube. NPS were performed according to the usual technique [4, 11]. Specimens were stored at 4°C after being collected.

### Nasal wash sampling

PCR tubes (CobasTM Roche Reference No. 07976577001-3) were filled with 1 ml of DMEM and 2ml of NaCl 0,9% was added in half of them. Consecutive NW and NPS specimens from 20 volunteers were obtained. NW was always performed first, and as follows: 3 ml of sterile saline solution was injected into the nostril using a 3 ml syringe and recovered into a plastic cup by leaning patients’ heads forward. Using the same syringe, a total 2 ml volume of the NW was transferred into a Cobas PCR tube containing 1 ml of DMEM media. NPS were performed according to the usual technique [4, 11]. They were then transferred into a Cobas PCR tube containing 1ml of DMEM media and 2ml of NaCl 0, 9% in order to compare the two techniques using equal media dilutions. Specimens were stored at 4°C after being collected.

*Video demonstrating the NW procedure available at: https://www.youtube.com/watch?v=3cMoR7hSPF8&feature=emb_title*

### SARS-CoV-2 RNA extraction and rRT-PCR

Viral RNA genome detection was performed by real-time RT-PCR using the Roche Cobas 6800 system (Cobas SARS-CoV-2 Ref 09175431190; Cobas SARS-CoV-2 Control kit Ref 09175440190; Cobas 6800/8800 Buffer Negative Control kit Ref 07002238190). This technology allows nucleic acid extraction, purification, PCR amplification and detection of SARS-CoV-2, targeting ORF1a/b and a pan-Sarbecovirus conserved region of the E protein gene.

### Dulbecco’s Modified Eagle Medium validation

We assessed the suitability of the Dulbecco’s Modified Eagle Medium (DMEM) for use in testing specimens by comparing it to the Universal Transport Medium (UTM). Ten positive NPS specimens were simultaneously spiked in 3ml DMEM and 3ml UTM and then analyzed by rRT-PCR using the Roche Cobas 6800 system.

### Statistical analyses

The correlation between the Ct values for ORF1 (arbitrary chosen for comparison) in NPS and in OPS or in NW specimens was evaluated using the Pearson correlation coefficient (*r*). The negative specimen by NW was arbitrarily assigned a Ct value of 45 and the two negative specimens by both OPS and NPS were excluded from the analysis. Correlation was also represented graphically using a simple linear regression *(figures 1 and 2)*. Statistical analyses were performed using IBM SPSS Statistics 25.

**Figure 1.**
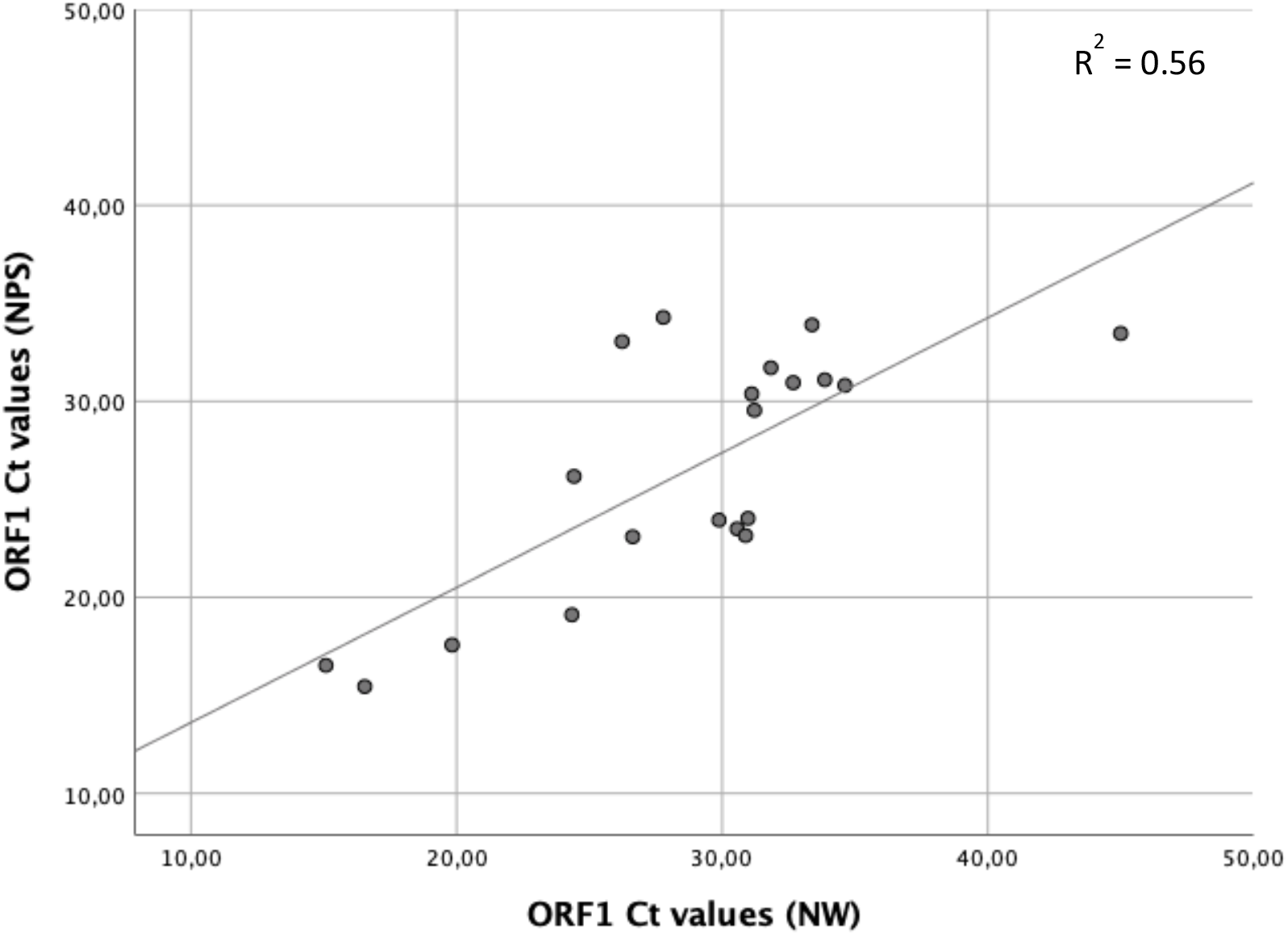
Correlation between rRT-PCR Cycle threshold (Ct) values obtained with nasal washes (NW) and with nasopharyngeal swabs (NPS). Each dot represents one of the 20 patients who had a NW and a NPS. One negative specimen by NW was arbitrary fixed at a Ct value of 45. The trend line is estimated by a simple linear regression, and the correlation coefficient R^2^ (square of the Pearson *r* value) is represented on the top right corner.

**Figure 2.**
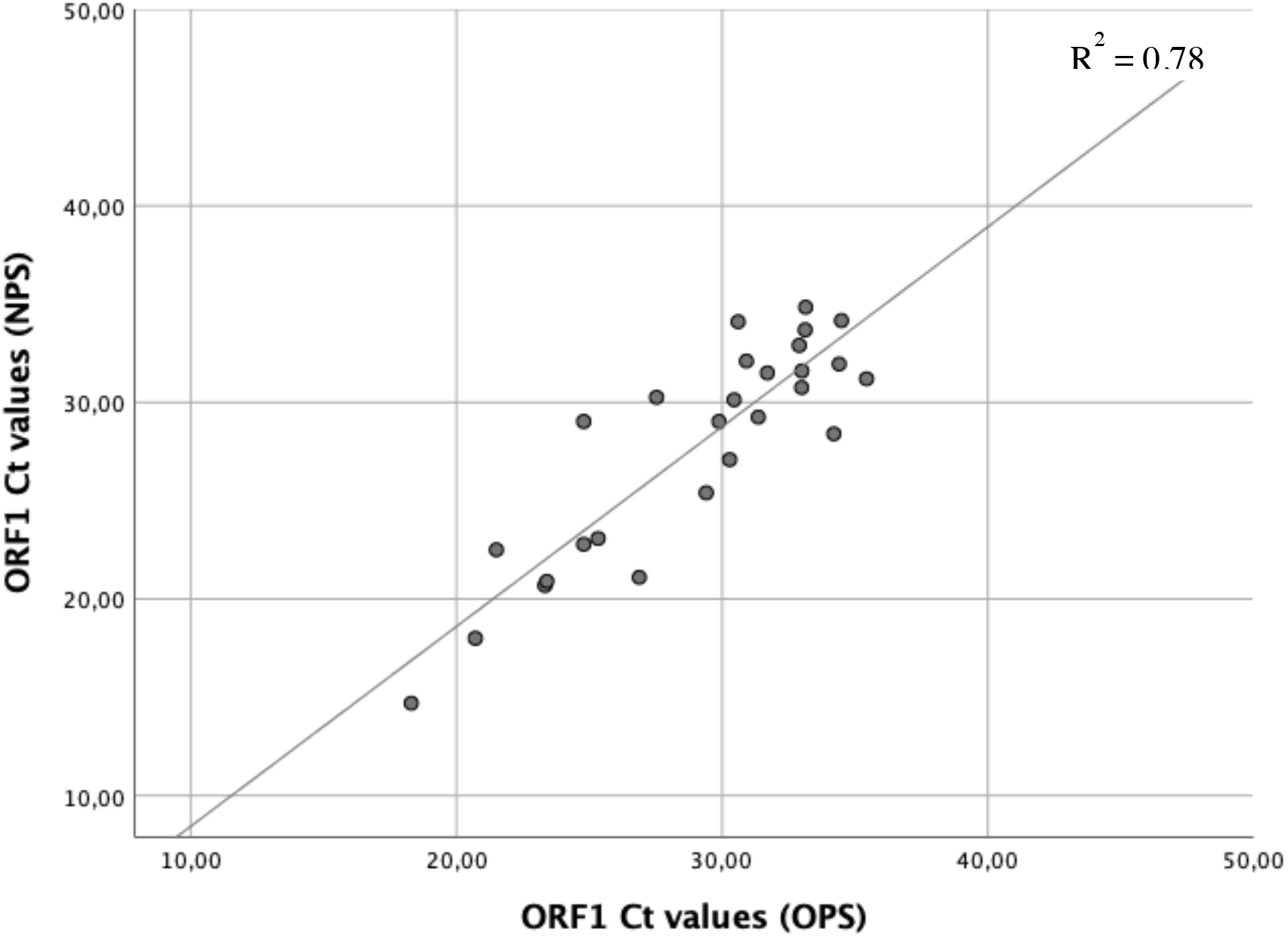
Correlation between rRT-PCR Cycle threshold (Ct) values obtained with oropharyngeal swabs (OPS) and with nasopharyngeal swabs (NPS). Each dot represents one of the 27 patients who had positive OPS and NPS. The trend line is estimated by a simple linear regression, and the correlation coefficient R^2^ (square of the Pearson *r* value) is represented on the top right corner.

## Results

### Dulbecco’s Modified Eagle Medium validation

The resulting cycle threshold (Ct) values were very similar when comparing the DMEM to the UTM. The delta Ct values ranged from 0.01 to 2.59 with a mean delta Ct value of 0.53 for ORF-1 and 0.67 for E-gene *(supplementary data, table 1)*.

### Sampling methods comparison

We compared the techniques in two groups of volunteers: the first group comprised 20 cases where a NW specimen was collected followed by a NPS specimen. The second group comprised 29 cases where an OPS specimen was collected followed by a NPS specimen. The clinical sensitivities of NW and OPS specimens were compared to those of the NPS specimens using the Ct values obtained by SARS-CoV-2 rRT-PCR *(supplementary data, table 2)*. Out of 20 cases, one patient that was positive with the NPS sampling with Ct values of 33.46 for ORF1 and 35.12 for the E-protein gene had negative results with the NW sampling. When comparing the NW sampling to NPS sampling, the mean delta Ct values were 1.77 (range -6.82 to 7.06) and 1.73 (range -7.79 to 8.25) for ORF1 and E-protein gene respectively *(supplementary data, table 2)*. The Pearson *r* was 0.75 (p<0.01), showing a statistically significant correlation between the ORF1 Ct values for the NPS and the NW specimens *(figure 1)*.

Out of 29 patients, two cases were negatives in both the OPS and the NPS specimens. When comparing OPS to NPS sampling, the mean delta Ct values were 1.24 (range -4.24 to 5.8) and 1.32 (range -4.63 to 7.6) for ORF1 and E-protein gene respectively *(supplementary data, table 2)*. The Pearson *r* was 0.88 (p<0.01), showing a statistically significant correlation between the ORF1 Ct values for the NPS and the OPS specimens *(figure 2)*.

## Discussion

Several studies have compared different types of upper respiratory tract specimens, and various collection methods have been compared to the gold standard method, the NPS [4, 11-13]. NW have also displayed promising upper respiratory virus detection rates [12, 14].

Regarding the NW samples, based on our results the clinical sensitivity seemed comparable to that of NPS specimens. A single NW sample was rRT-PCR negative, whereas the NPS one collected consecutively from the same patient was positive. However the high Ct values of these samples suggest that the viral RNA present in both specimens from this volunteer was close to the limit of detection, and we cannot affirm that the clinical sensitivity of NW is below that of NPS specimens for SARS-CoV-2 RNA detection based on this single observation. On the quantitative level, the mean delta Ct values seemed acceptable and the correlation between NW and NPS was reinforced by the statistical analyses.

Our results also indicate a comparable clinical sensitivity between OPS and NPS at the qualitative level, since all patients with positive SARS-CoV-2 rRT-PCR results from NPS specimens, even those with high Ct values, also tested positive by OPS. Regarding the analytical sensitivity at the quantitative level, we obtained a significant correlation between OPS and NPS specimens.

Concerning the transport medium, our results suggest that the DMEM seems to be suitable for SARS-CoV-2 detection.

On the practical side, OPS and NW appeared to be better tolerated by patients, although this needs to be confirmed by using appropriate patient scoring. Another practical advantage of OPS over NPS in an equipment shortage setting is that adequate rigid swabs are much more readily available than those used for NPS. Nasal washes present a valuable advantage as they can be performed without the need of specific swabs and with a minimal use of tools that are unlikely to be in shortage anyway. Moreover, both procedures seemed to cause less coughing than the NPS procedure, which represents a major advantage when considering the exposure of healthcare workers to SARS-CoV-2.

Limitations to our study include the relatively small sample size, and further evaluation would be needed to reach a definitive conclusion. The statistical analyses were also underpowered and should be interpreted with caution.

In conclusion, OPS and NW seem to be reliable alternative upper respiratory tract sampling methods for the molecular detection of SARS-CoV-2, and DMEM can be used as an alternative to commercial UTM, particularly in crisis settings. Further studies with a higher number of samples would still be needed to firmly conclude to an equivalence between these sampling methods. Nevertheless, the increased testing versatility offered by these substitutes should be greatly welcomed in the COVID-19 global crisis setting.

## Data Availability

The authors confirm that the data supporting the findings of this study are available within the article and its supplementary material.

## Acknowledgments

We would like to thank Benedikt Huttner for his help concerning statistical analyses, and Erik Boehm for his help in writing and correcting the manuscript.

## Compliance with Ethical Standards

### Research involving human participants

According to the Swiss Ethics Committees on Research Involving Humans standards, this study took place in the context of a method quality validation in an emergency setting, and therefore did not require any authorization by our ethic committee.

### Consent to participate

Oral informed consent was obtained from all individual participants included in the study.

## Supplementary data: tables

**Supplementary table 1.** Comparison of DMEM and UTM media. The delta Ct value for target ORF 1 between the two transport media containing the same specimen ranged between 0.01 and 2.59. One specimen turned out negative in both media. Two specimens were very close to the detection limit and yielded undetectable results for one or more targets (patient a: ND/ND and 36.43/ND, patient e: ND/37.81 and 36.3/37.82). ND: not detected. NA: not applicable.

| Patient | DMEM |  | UTM |  | ΔCt values |  |
| --- | --- | --- | --- | --- | --- | --- |
|  | ORF 1 | E-gene | ORF 1 | E-gene | ORF 1 | E-gene |
| a | ND | ND | 36.43 | ND | NA | NA |
| b | 28.8 | 29.39 | 29.03 | 30.08 | 0.23 | 0.69 |
| c | ND | ND | ND | ND | NA | NA |
| d | 31.17 | 32.55 | 28.89 | 29.96 | 2.81 | 2.59 |
| e | ND | 37.81 | 36.3 | 37.82 | NA | 0.01 |
| f | 24.95 | 25.43 | 25.01 | 25.11 | 0.06 | 0.32 |
| g | 20.87 | 21.6 | 20.84 | 20.94 | 0.03 | 0.66 |
| h | 32.22 | 33.64 | 31.81 | 32.73 | 0.41 | 0.91 |
| i | 28.8 | 29.51 | 28.9 | 29.41 | 0.1 | 0.1 |
| j | 29.25 | 30.14 | 29.17 | 30.04 | 0.08 | 0.1 |

**Table 2.** Sampling methods comparison. **a**. Ct values of NP swab specimens compared to Ct values of NW specimens by RT-PCR in 20 patients. The mean delta Ct value shows a slightly better overall sensitivity with NP swabs (average 1.7). Patient 15’s NW sample was negative despite a positive NP specimen. **b**. Ct values of NP swab specimens compared to Ct Values of OP swab specimens in RT-PCR in 29 patients. Patients 1 and 28 were negative both in NP and OP swabs. The mean delta Ct value shows a slightly overall better sensitivity with NP swabs (average 1.2-1.3). The specimen collection was done at the same time.

| a. | NP swab + 1ml DMEM + 2ml NaCl |  | 2ml NASAL WASH + 1ml DMEM |  | delta CT |  |
| --- | --- | --- | --- | --- | --- | --- |
| patients | ORF1 Ct value | E-gene Ct value | ORF1 Ct value | E-gene Ct value | ORF1 | E-gene |
| 1 | 23.5 | 23.9 | 30.56 | 31.21 | 7.06 | 7.31 |
| 2 | 34.28 | 35.33 | 27.78 | 28.67 | -6.5 | -6.66 |
| 3 | 23.15 | 23.63 | 30.88 | 31.88 | 7.73 | 8.25 |
| 4 | 30.95 | 32.84 | 32.67 | 33.95 | 1.72 | 1.11 |
| 5 | 24.03 | 24.06 | 30.97 | 31.51 | 6.94 | 7.45 |
| 6 | 23.94 | 24.32 | 29.88 | 30.93 | 5.94 | 6.61 |
| 7 | 30.38 | 31.44 | 31.11 | 31.71 | 0.73 | 0.27 |
| 8 | 19.11 | 19.43 | 24.34 | 24.65 | 5.23 | 5.22 |
| 9 | 29.54 | 30.64 | 31.21 | 32.34 | 1.67 | 1.7 |
| 10 | 15.45 | 15.71 | 16.54 | 16.64 | 1.09 | 0.93 |
| 11 | 31.1 | 32.28 | 33.86 | 35.5 | 2.76 | 3.22 |
| 12 | 31.71 | 33.79 | 31.83 | 32.8 | 0.12 | -0.99 |
| 13 | 33.05 | 34.48 | 26.23 | 26.69 | -6.82 | -7.79 |
| 14 | 16.53 | 16.69 | 15.08 | 15.32 | -1.45 | -1.37 |
| 15 | 33.46 | 35.12 | NEG | NEG | NA | NA |
| 16 | 26.17 | 26.71 | 24.42 | 24.47 | -1.75 | -2.24 |
| 17 | 17.57 | 18.01 | 19.83 | 20.12 | 2.26 | 2.11 |
| 18 | 23.09 | 23.9 | 26.63 | 28.32 | 3.54 | 4.42 |
| 19 | 30.81 | 32.7 | 34.62 | 37.18 | 3.81 | 4.48 |
| 20 | 33.9 | 35.96 | 33.38 | 34.81 | -0.52 | -1.15 |
| Mean delta CT |  |  |  |  | 1.76631579 | 1.73052632 |
| b. | NP swab + 3ml DMEM |  | OP swab + 3ml DMEM |  | Delta Ct values |  |
| patients | ORF1 Ct value | E-gene Ct value | ORF1 Ct value | E-gene Ct value | ORF1 | E-gene |
| 21 | NEG | NEG | NEG | NEG | NA | NA |
| 22 | 28.4 | 29 | 34.2 | 36.6 | 5.8 | 7.6 |
| 23 | 21.1 | 22 | 26.88 | 27.67 | 5.78 | 5.67 |
| 24 | 31.2 | 32.47 | 35.43 | 37.83 | 4.23 | 5.36 |
| 25 | 25.4 | 26.3 | 29.4 | 30.3 | 4 | 4 |
| 26 | 14.7 | 15.8 | 18.3 | 18.7 | 3.6 | 2.9 |
| 27 | 27.09 | 27.5 | 30.28 | 30.73 | 3.19 | 3.23 |
| 28 | 18 | 18.38 | 20.71 | 21.14 | 2.71 | 2.76 |
| 29 | 20.69 | 21.34 | 23.33 | 23.56 | 2.64 | 2.22 |
| 30 | 20.9 | 21.6 | 23.4 | 23.9 | 2.5 | 2.3 |
| 31 | 31.95 | 34.17 | 34.4 | 36.9 | 2.45 | 2.73 |
| 32 | 23.08 | 23.61 | 25.33 | 25.72 | 2.25 | 2.11 |
| 33 | 30.75 | 32 | 32.99 | 33.85 | 2.24 | 1.85 |
| 34 | 29.25 | 30.13 | 31.36 | 32.62 | 2.11 | 2.49 |
| 35 | 22.78 | 23.45 | 24.79 | 25.24 | 2.01 | 1.79 |
| 36 | 31.6 | 32.5 | 33 | 34 | 1.4 | 1.5 |
| 37 | 29.03 | 29.97 | 29.88 | 30.69 | 0.85 | 0.72 |
| 38 | 34.16 | 36.8 | 34.49 | 35.96 | 0.33 | -0.84 |
| 39 | 30.13 | 31.57 | 30.45 | 31.28 | 0.32 | -0.29 |
| 40 | 31.5 | 34 | 31.7 | 33.6 | 0.2 | -0.4 |
| 41 | 32.9 | 35.1 | 32.9 | 34.8 | 0 | -0.3 |
| 42 | 33.69 | 36.5 | 33.12 | 34.43 | -0.57 | -2.07 |
| 43 | 22.5 | 23.00 | 21.5 | 22.30 | -1 | -0.7 |
| 44 | 32.1 | 33.56 | 30.91 | 32.96 | -1.19 | -0.6 |
| 45 | 30.26 | 31.01 | 27.53 | 28.13 | -2.73 | -2.88 |
| 46 | 34.1 | 36.6 | 30.6 | 32.3 | -3.5 | -4.3 |
| 47 | 29.03 | 30.05 | 24.79 | 25.42 | -4.24 | -4.63 |
| 48 | NEG | NEG | NEG | NEG | NA | NA |
| 49 | 34.84 | 36.83 | 33.14 | 35.2 | -1.7 | -1.63 |
| Mean delta CT |  |  |  |  | 1.2474074<br>1 | 1.1329629<br>6 |

